# Machine-learning-based analysis of transcriptomics data for the identification of molecular signatures in cancer

**DOI:** 10.1101/2025.10.09.25337647

**Authors:** Kulandai Arockia Rajesh Packiam, Sharmistha Majumdar, Anirban Dasgupta

## Abstract

Early detection and treatment of head and neck squamous cell carcinoma (HNSC) and oral squamous cell carcinoma (OSCC) could decrease the existing high mortality rates due to these cancers. The need for biomarker identification is crucial to detect HNSC and OSCC in their initial stages enabling prompt treatment. The present study establishes a machine learning-based framework employing a two-step feature selection strategy consisting of analysis of variance and coupled support vector machines - recursive feature elimination that delineates biomolecular signatures in HNSC and OSCC. Based on these signatures, classification models with high classification and prediction accuracies were developed which highlighted the significance of selected 22 HNSC and 23 OSCC candidate genes. Further, K-means clustering supported this finding by displaying a clear demarcation of the normal and tumor classes while previous literature confirmed the importance of the biomolecular signatures in several cancers. Specifically, it has been reported that FAM107A, FAM3D and CXCR2 could be probable diagnostic or prognostic candidates while RRAGD, PPL, AQP7, SORT1, MAB21L4 and UBL3 were earlier suggested to have therapeutic roles in cancer. Three overlapping genes namely, ENDOU, RRAGD and SMIM5 could be imperative because of their commonality between HNSC and OSCC. Likewise, seven features related to plasma membrane i.e., CEACAM1, COBL, GPX3, HCG22, MUC21, PAX9 and SMIM5 were identified that could essentially be targeted as non-invasive diagnostic, prognostic or therapeutic candidates. Therefore, the HNSC and OSCC biomolecular signatures obtained as a result of the present computational approach showed promising potentials as a diagnostic or prognostic or therapeutic biomarker.

## Introduction

Cancer, defined by uncontrolled growth of cells leading to adjacent tissue invasion and metastasis (Jiang is one of the prime causes for most numbers of deaths counting as almost one in six deaths, globally (WHO Fact sheet: Cancer). Specifically, head and neck squamous cell carcinoma (HNSC) and oral squamous cell carcinoma (OSCC) has posed a serious health concern worldwide (Borse et al. 2020; Ye et al. 2022). HNSCs are tumors that lie on epithelial tissues of upper digestive tract and within HNSC, cancerous growth in the mouthparts namely, lips, cheeks, sinuses, tongue, hard and soft palate, the base of the mouth extended to the oropharynx is referred to as OSCC. Most commonly, HNSCs and OSCCs are caused due to heavy tobacco consumption that includes either smoking or chewing tobacco. Apart from the excess alcohol consumption, poor oral hygiene or viral infections are other minor causes for HNSC and OSCC (Tanriver et al. 2021; Kozłowska et al. 2021). Generally, HNSCs include conventional treatment strategies such as surgery, chemotherapy and radiation which is administered as a single or multi-modal therapy. However, even after such intense and systematic interventions, these tumors usually have high chances of recurrence due to acquired immunity (Kozłowska et al. 2021). Additionally, lower survival rates in HNSC is partially due to the diagnosis at a later stage and hence, early diagnosis can improve the HNSC survival rates (Economopoulou et al. 2019). Similarly, surgery is performed to treat OSCCs and has high success rates but most of the cases are diagnosed very late in their advanced stages, resulting in higher mortalities (Alabi et al. 2021; Tanriver et al. 2021). Early diagnosis of OSCC and a clarity on its prognosis can remarkably reduce the high mortality rates due to OSCCs. But the cancerous lesions are completely asymptomatic, too small in size, harmless and painless during the early stages of OSCC, so that a prompt diagnosis upon examination is not often successful. Novel molecular signatures with respect to HNSCs and OSCCs may be helpful in identifying novel candidates for the early diagnosis and subsequent treatment strategies for these cancer sub-types.

Cancer, in general, is a multi-layered disease involving complex interactions at various levels within cells, tissues of an organism (Paul 2020). Within a cell, up or down-regulation of biomolecules such as genes, proteins or metabolites may occur at different levels such as the genetic, proteomic, or metabolomic level caused by genetic mutation or other environmental factors (Cho 2010). The identification and detailed study of such biomolecules can lead to deeper insights into cancer diagnostics and treatment. With the recent advancement in molecular profiling technology, volumes of multi-omics data, specifically, transcriptomics or RNA sequencing (RNA-seq) data for several cancer-subtypes are now publicly available. Traditional approaches to analyse RNA-seq data such as differentially expressed genes (DEGs) analysis successfully identifies the up- and down-regulated genes that play key roles in cancer. Recently, machine learning (ML), a sub-field of artificial intelligence aiming at data-driven modelling to simplify the data analysis by developing prediction models, has been used in omics data analysis (Tran et al. 2021). By means of ML-based approaches, an apparent classification of tumor cases against normal cases for HNSC and OSCC is possible based on the significant features. These significant features can act as important leads towards identification of biomarkers precisely, diagnostic, prognostic or therapeutic candidates. Since biomarker identification remains crucial in case of HNSC and OSCC, the availability of a large RNA-seq dataset will be helpful in establishing the ML-based data analysis pipeline to select important biomolecular signatures as biomarkers. At the same time, the existing literature-based findings will also allow subsequent validation of the importance of ML-based predicted biomolecular signatures.

The present study exhibits a simplified and an effective protocol for the identification of significant biomolecular signatures in HNSC and OSCC by applying machine learning (ML) approaches. Briefly, a pipeline was developed that constituted of a two-step feature selection strategy followed by model training and testing with various classifiers to classify the given samples into one of the two classes, i.e., tumor and normal. ANOVA was used to select top 100 features which were then, further reduced to significant numbers of features using a coupled support vector machine (SVM) – recursive feature elimination (RFE) method. The significant features altogether form the biomolecular signature. The models were trained using data pertaining to biomolecular signature and showed high performance metrics which readily suggests that the selected features are of paramount importance in HNSC and OSCC. Additionally, the significance of selected biomolecular signatures corresponding to HNSC and OSCC was further confirmed by unsupervised clustering that yielded a clear demarcation with two clusters each for normal and tumor samples. To confirm further whether the identified HNSC or OSCC features play prominent functions in respective cancers, a literature-based validation was performed. Previous studies revealed that most of the selected features from HNSC and OSCC biomolecular signatures are known to have prognostic or therapeutic functions or are significantly expressed in differential levels in cancer than normal. In contrast, the rest of the features remain unexplored, thereby necessitating the need for further investigation which may be fruitful in identifying novel biomarkers or therapeutic agents.

## Materials and methods

### Software, programming languages and packages employed

The programming language, R v. 4.2.1 was employed to download the datasets using the Bioconductor package while the complete machine learning pipeline has been implemented in Python v. 3.10.4 using Jupyter notebook v. 6.4.12 and scikit learn library. Other key Python packages used were shap, seaborn, matplotlib, numpy and pandas.

### Dataset

The transcriptomics data consisting of 60,660 transcripts including both coding and non-coding genes for 713 samples. The samples corresponded to two TCGA projects, namely, TCGA-HNSC (n = 548) and CPTAC3-HNSC (n = 165) and were downloaded from the Genomic Data Commons (GDC) Data Portal (https://portal.gdc.cancer.gov/) (Grossman et al. 2016). The dataset represented the gene expression (RNA-seq) data in Fragments Per Kilobase of transcript per Million mapped reads upper quartile (FPKM-UQ) units on the platform— the Illumina Human Methylation 450K platform. From the HNSC samples, 442 OSCC samples were selected based on the primary sites such as the lip, the base of the tongue, gum, tonsil, palate, the floor of the mouth, other and unspecified parts of the mouth, other and unspecified parts of tongue, other and ill-defined sites in lip and oral cavity and pharynx (Uddin et al. 2022). For simplicity, the transcriptomics or T-omics data corresponding to HNSC and OSCC used in the current research are henceforth, called as HNSC T-omics and OSCC T-omics data, respectively.

### Feature selection methods

Analysis of variance (ANOVA) was applied as the first step of the feature selection strategy in the present study. ANOVA is a filter-based method where the means of the different groups are statistically compared for equality using F-tests. Higher F-scores indicate that the feature has higher inter-group dispersion resulting in a clear separation within the means (Gomes et al. 2022). Therefore, top 100 features were chosen from the initial 60,660 features by employing ANOVA F-tests in Python using the SelectKBest algorithm from scikit_learn with score_func = f_classif and k=100. The top 100 features from HNSC and OSCC were further screened for most significant features using recursive feature elimination (RFE). Unlike ANOVA, RFE is a wrapper-based method which completely relies on a classifier and/or regressor to perform the corresponding classification or regression task. In RFE, features are ranked and those with lowest importance are removed iteratively until certain criteria, such as number of minimum features retained, are achieved (Wang et al. 2022). RFE was performed using RFECV with a stratified 5-fold cross validation, minimum features to be selected as 20 and weighted F1 score as a scoring function on a “linear kernel” of support vector machine (SVM) classifier with default parameters. To check the reproducibility of SVM-RFE, the shuffle parameter during StratifiedKFold was set to “TRUE” and the random state of the seed was changed every time for five repeats. Additionally, the feature importance was computed based on SHapley Additive exPlanations (SHAP) values to verify the significance of the selected genes. SHAP implemented in the shap library of Python was used for computing feature importance (Supplementary information). T-omics data which was trimmed based on Top 100 features and biomarker signatures are denoted as Top 100 data and BioSign data, respectively. Similarly, T-omics data corresponding to HNSC and OSCC biomolecular signatures are termed HNSC and OSCC BioSign data respectively.

### Classification models and performance metrics

ML models were trained using different classifiers namely logistic regression (LogR), random forest (RF), gradient boosting (GB), eXtreme Gradient Boosting or XGBoost (XGB), multilayer perceptron (MLP) and SVM by applying 5-fold cross validation (CV). In 5-fold CV, the training datasets are randomly split into five smaller subsets, wherein four folds are used for the training while the fifth fold is used as the internal model testing dataset and the whole process is iterated five times until all the five folds have been used as testing datasets. The performances of the classifiers were measured as accuracy, precision, recall and F1 scores and reported as the average of all the performances of all the five subsets.

### Clustering tasks

Dimensionality reduction and cluster visualization in a 2-dimentional space was done using Principal components analysis (PCA) with n_components = 2 on Top-100 and BioSign data for both HNSC and OSCC. Subsequently, unsupervised clustering was performed by using KMeans clustering with n_clusters = 2, init=“k-means++”, n_init=10, and max_iter=100 as the parameters. Silhouette coefficient (SC), adjusted random index (ARI) and percentage of correctly clustered samples or correctly clustered index (CCI) were used to evaluate the performance of the clustering tasks. The function, silhouette_score was used to estimate SC that ranges between −1.0 to 1.0 where a higher value represents a higher degree of separation during clustering (Owens et al. 2021; Mahalanabis et al. 2022). ARI denotes a chance-adjusted measure of similarity or the agreement between the clustering of the predicted class to the actual class (Mahalanabis et al. 2022) and was determined using the function, adjusted_rand_score. The values of ARI vary from −0.5 to 1 depending on the similarities within clusters and the higher ARIs indicate strong agreement between clusters. CCI was computed as the percentage of true clustering achieved, i.e., the ratio of the samples that are correctly clustered in both the clusters to the total number of samples that were used for clustering. All the functions were implemented using Scikit-learn Python library until otherwise mentioned.

## Results

### ML-based workflow

An ML pipeline was developed to identify the biomolecular signatures in HNSC and OSCC, and the features from BioSign genes were further used in the classification tasks to demarcate the tumor and normal cases within the two cancer types (Figure 1). T-omics data were extracted from the data repository TCGA considering the two projects, TCGA-HNSC and CPTAC3-HNSC which constituted 82% and 18% of the total cases, respectively (Figure S1). Additionally, male population comprised three-fourth of the total cases while almost the same proportions represented the cases where patients were of fifty-five years of age and above. Out of the 713 HNSC T-omics data, 611 corresponded to Tumor samples while 102 for Normal samples. 62% of the total HNSC cases were sub-categorised as “OSCC” by using the primary location of the tumor. The HNSC and OSCC T-omics were randomly split into 80% training datasets that were used for further feature selection and model training. And the remaining 20% of the total cases in both HNSC and OSCC were withheld as independent datasets for validation. ANOVA was used as the first step of feature reduction to select statistically significant features and top 100 features were chosen from the total of 60,660 initial features. SVM-RFE was subsequently employed in the second step of feature selection to determine the biomolecular signature for HNSC and OSCC from the top 100 features. Feature selection using SVM-RFE was optimized for five runs and the key genes commonly selected in all the runs were chosen (Figure S2 and S3). After each feature selection step, i.e., ANOVA and SVM-RFE, respectively, models were trained using the training datasets, Top 100 and BioSign data, for both HNSC and OSCC. The models were validated by using the independent test datasets while unsupervised clustering on the training datasets followed by literature-based confirmation was performed to validate the importance of selected candidates for biomolecular signatures.

**Figure 1.**
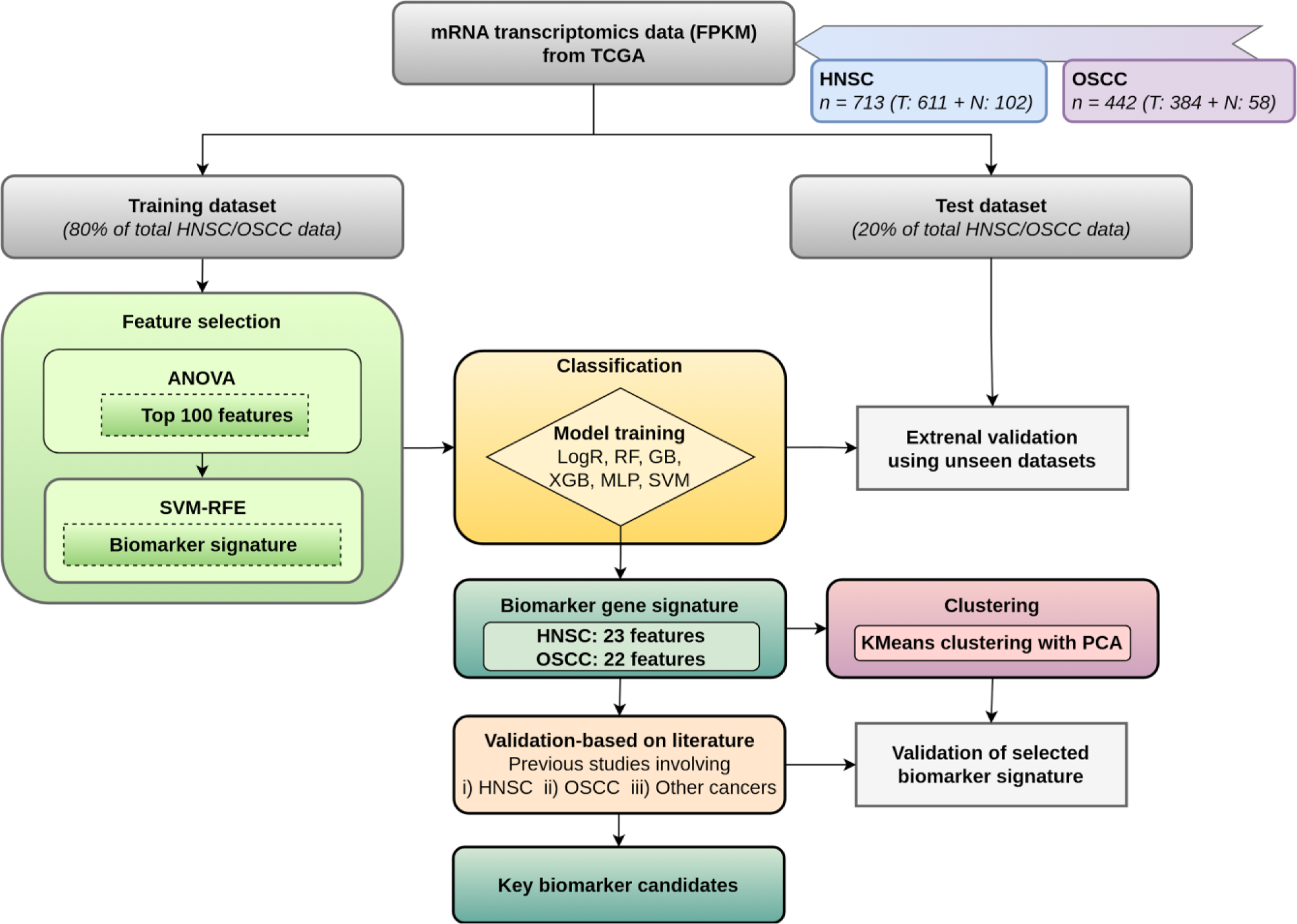
Machine learning pipeline for identifying biomolecular signatures in HNSC and OSCC. Legends: Head and neck squamous cell carcinoma (HNSC), Oral squamous cell carcinoma (OSCC), Logistic Regression (LogR), Random Forest (RF), Gradient Boosting (GB), XGBoost (XGB), Multi-Layer Perceptron (MLP), Support vector machine (SVM), and Recursive feature elimination (RFE).

### Identification of biomolecular signature in HNSC and OSCC using ANOVA and SVM-RFE

ANOVA, the first step of feature selection, has been the most popular as well as the most effective method for dimensionality reduction for omics data (Kirpich et al. 2018). Our preliminary feature selection based on ANOVA yielded top 100 features in HNSC (Table S1) and OSCC (Table S2). A previous study dealing with the DEGs in HNSC (Shaikh et al. 2019) confirmed the significance of the ANOVA-based selection of top 100 features. Out of the 100 top features from ANOVA, 90 features in HNSC and 93 features in OSCC have been detected as DEGs in the study by Shaikh et al. 2019. This clearly shows that the selected top 100 features are statistically significant and also, ruled out the necessity to perform an independent DEG analysis for OSCC and HNSC to validate the significance of the selected features. We, then, applied the second feature selection strategy, SVM-RFE, to further narrow down the top 100 features to a smaller subset of biomolecular signatures in both HNSC and OSCC. SVM-RFE was successfully implemented as a feature selection method for a wide number of omics studies specifically in various cancer sub-types to date (Lin et al. 2018). Our results also showed that by employing SVM-RFE as the second step of feature selection, a total of 22 and 23 features, respectively, were identified as biomolecular signatures (Figure 2) from the top 100 ANOVA-based features for HNSC and OSCC. The gene distribution and expression plots for the selected 22 HNSC (Figures S4 and S6) and 23 OSCC features (Figures S5 and S7) also supported the notion that the selected features are differentially expressed in normal and tumor samples of the HNSC and OSCC sub-types. Furthermore, all the selected features in the biomolecular signature are noticed to be down-regulated in both the HNSC and OSCC cancer sub-types.

**Figure 2.**
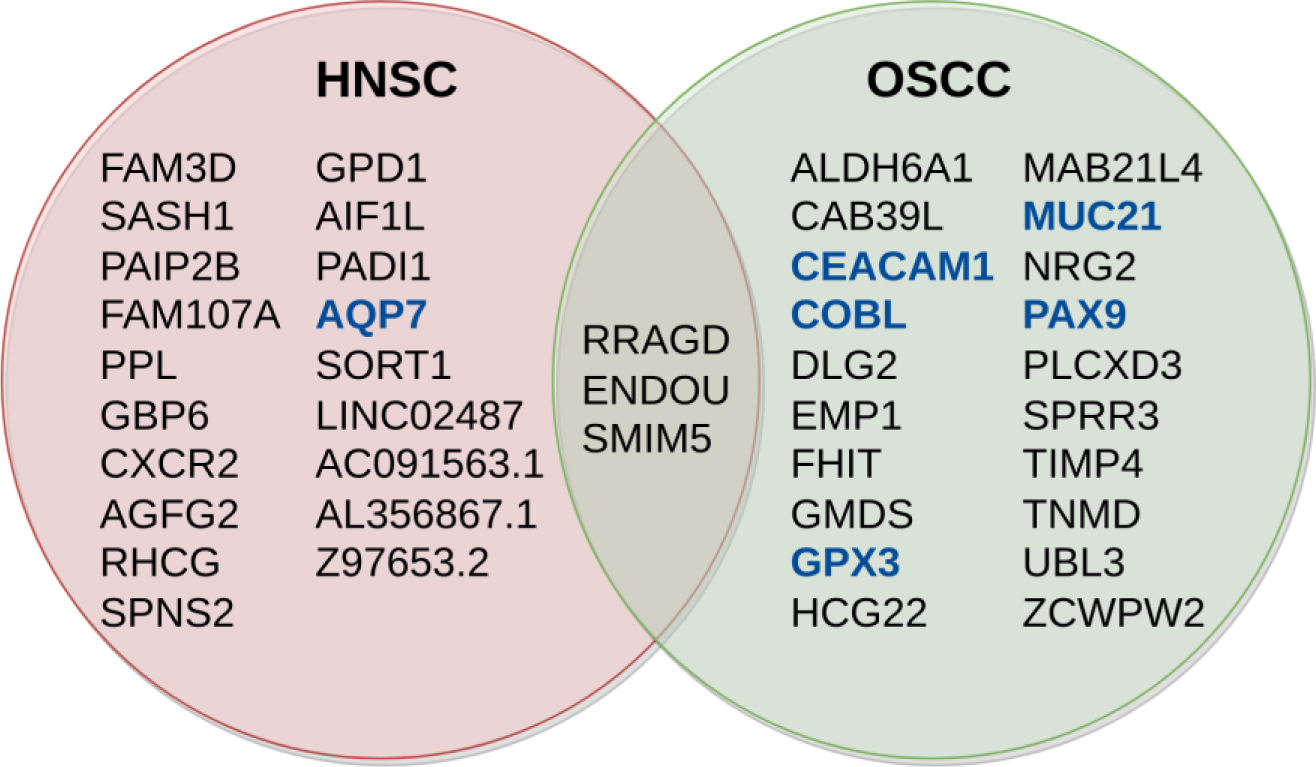
Selected features in the biomolecular signature for HNSC and OSCC. Intersection represents the genes that are common between HNSC and OSCC while the genes associated with cellular or plasma membranes are shown in blue and bold.

### Performance of various classification models with top 100 features and biomolecular signature

Conceptually, it is hypothesized that the selected top 100 features and the biomolecular signatures will be able to perform the classification of the tumor or normal samples with a greater accuracy. To determine the importance of the selected top 100 features and the biomolecular signatures towards classification, ML models were trained using six different classifiers such as Logistic Regression (LogR), Random Forest (RF), Gradient Boosting (GB), XGBoost (XGB), Multi-Layer Perceptron (MLP), and Support vector machine (SVM). The performances of the respective classifiers were benchmarked by training models on these classifiers using datasets based on the top 100 features and biomolecular signature for both cancer sub-types, HNSC and OSCC, followed by independent testing to validate the classification models. An overall average accuracy of 0.95 to 0.99 was achieved in case of both HNSC and OSCC when the classification models employing different classifiers were trained on datasets pertaining to both top 100 features and biomolecular signature with five-fold cross validation (Table 1). No drastic change in the classification accuracy was seen when the models trained on biomolecular signature from those trained with top 100 features as features for both HNSC or OSCC. This established the fact that the biomolecular signature for both HNSC or OSCC can efficiently perform the classification task on a par with top 100 features. Secondly, it also suggested that the biomolecular signature classifies normal and tumor cases accurately regardless of the classifier-type employed for the classification tasks.

**Table 1.**
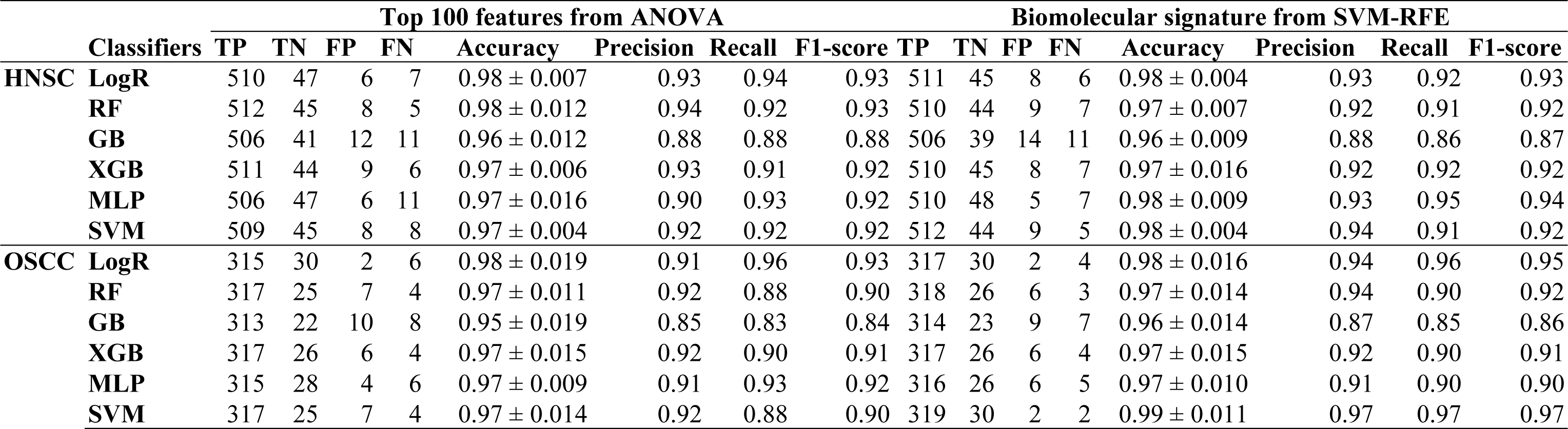
Benchmarking different classifiers during the training of classification tasks.

In addition to achieving high accuracy, it is crucial for the classification model to attain maximal numbers of correctly predicted tumor instances i.e., “true positive” or TP and minimal numbers of wrongly predicted normal instances, i.e., “false negatives” or FN. In other words, the classification models should be able to predict most of the tumors as tumors while keeping the incorrect predictions of tumors as normal to a minimum. During the classification of HNSC normal and tumor, LogR, MLP and SVM showed a slight increase in accuracies and TPs and decrease in FNs when trained on BioSign data compared to Top 100 data. The classification model using GB displayed no change in performance while ML models employing RF and XGB resulted in a small decrease in classification performances upon training using BioSign data against top 100 data. Similarly, in case of OSCC, all the classifiers except XGB gave an improved performance for models trained on BioSign data than top 100 data whilst no notable change was observed in the performance of XGB for models trained on the two datasets. On the whole, it was noticed that SVM tends to outperform other classifiers in terms of its performance on BioSign data in case of both cancers, HNSC and OSCC.

The receiver operating characteristic (ROC) curves were plotted for the class tumor for all the six classification models based on Top 100 and BioSign data for both HNSC and OSCC (Figure 3). Alternatively, the significance of the biomolecular signatures of HNSC and OSCC on different classifiers were plotted using SHAP values (Tables S8 and S9) which suggested that the feature importance varies with respect to the classifier. Therefore, these results suggested that by using the biomolecular signature as features, most of the classifiers and specifically, SVM, can perform classification of the tumor and normal cases accurately with high numbers of predicted TPs and low FNs.

**Figure 3.**
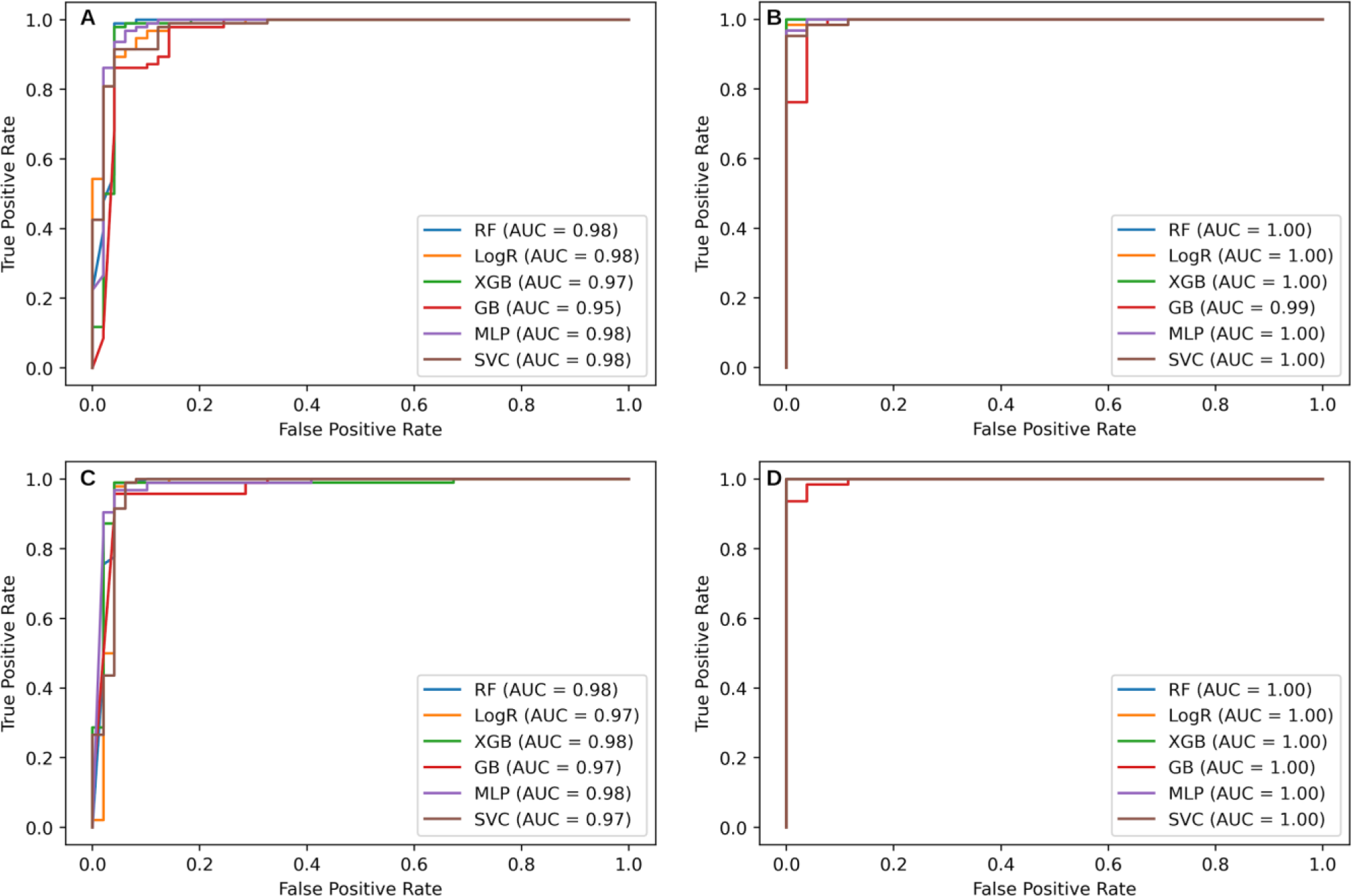
ROC curves for various classifiers corresponding to top 100 features in (A) HNSC and (B) OSCC, and molecular signatures in (C) HNSC and (D) OSCC.

**Figure 4.**
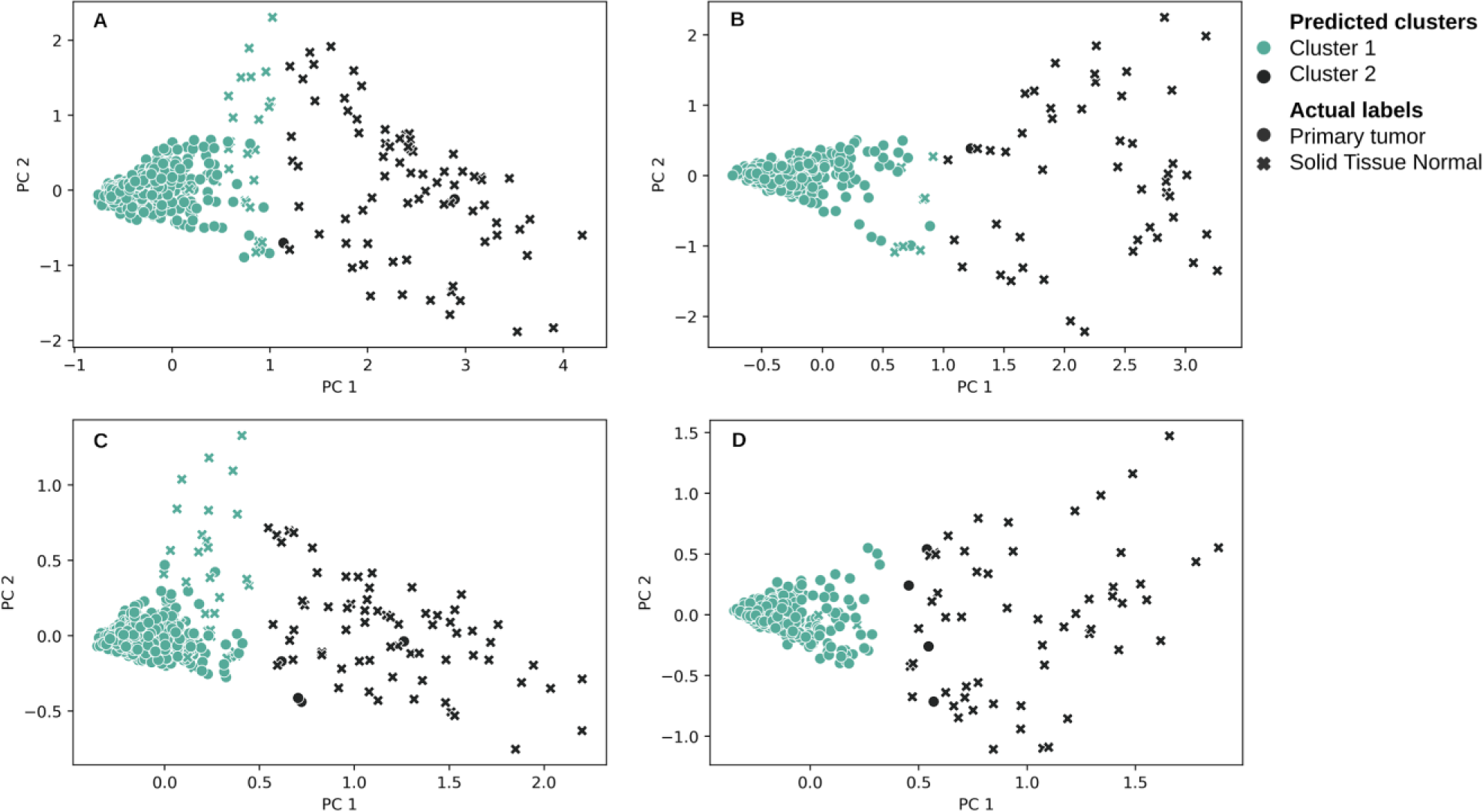
Clustering of top 100 features in (A) HNSC and (B) OSCC, and molecular signatures in (C) HNSC and (D) OSCC.

### Validation of the classification models using independent testing

The trained models were tested on independent unseen datasets to validate the importance of selected features within biomolecular signatures. Specifically, SVM gave a high prediction accuracy of 0.95 along with highest numbers of TPs (94) and nil FNs for models trained on HNSC BioSign data while the other three classifiers namely, RF, XGB and MLP gave a prediction accuracy of 1 with 63 TPs and nil FNs when tested on models trained with OSCC BioSigndata. On the whole, the prediction accuracies of the classification models trained on the BioSign were found to be elevated than those trained on Top 100 data for both HNSC and OSCC (Table 2). Therefore, independent testing supports the notion that the selected HNSC and OSCC biomolecular signatures are significant and that the models trained on various classifiers using BioSign data can perform better than models trained on Top 100 data.

**Table 2.** Validation of various classification models via independent testing.

|  | Classifiers | Top 100 features from ANOVA |  |  |  |  |  |  |  | Biomolecular signature from SVM-RFE |  |  |  |  |  |  |  |
| --- | --- | --- | --- | --- | --- | --- | --- | --- | --- | --- | --- | --- | --- | --- | --- | --- | --- |
|  |  | TP | TN | FP | FN | Accuracy | Precision | Recall | F1-score | TP | TN | FP | FN | Accuracy | Precision | Recall | F1-score |
| **HNSC** | **LogR** | 93 | 40 | 9 | 1 | 0.93 | 0.94 | 0.90 | 0.92 | 93 | 42 | 7 | 1 | 0.94 | 0.95 | 0.92 | 0.94 |
|  | **RF** | 93 | 45 | 4 | 1 | 0.97 | 0.97 | 0.95 | 0.96 | 93 | 46 | 3 | 1 | 0.97 | 0.97 | 0.96 | 0.97 |
|  | **GB** | 89 | 42 | 7 | 5 | 0.92 | 0.91 | 0.90 | 0.91 | 90 | 44 | 5 | 4 | 0.94 | 0.93 | 0.93 | 0.93 |
|  | **XGB** | 93 | 43 | 6 | 1 | 0.95 | 0.96 | 0.93 | 0.94 | 93 | 45 | 4 | 1 | 0.97 | 0.97 | 0.95 | 0.96 |
|  | **MLP** | 92 | 44 | 5 | 2 | 0.95 | 0.95 | 0.94 | 0.94 | 93 | 44 | 5 | 1 | 0.96 | 0.96 | 0.94 | 0.95 |
|  | **SVM** | 93 | 36 | 13 | 1 | 0.90 | 0.93 | 0.86 | 0.88 | 94 | 42 | 7 | 0 | 0.95 | 0.97 | 0.93 | 0.94 |
| **OSCC** | **LogR** | 60 | 26 | 0 | 3 | 0.97 | 0.95 | 0.98 | 0.96 | 62 | 26 | 0 | 1 | 0.99 | 0.98 | 0.99 | 0.99 |
|  | **RF** | 63 | 26 | 0 | 0 | 1.00 | 1.00 | 1.00 | 1.00 | 63 | 26 | 0 | 0 | 1.00 | 1.00 | 1.00 | 1.00 |
|  | **GB** | 59 | 25 | 1 | 4 | 0.94 | 0.92 | 0.95 | 0.93 | 62 | 25 | 1 | 1 | 0.98 | 0.97 | 0.97 | 0.97 |
|  | **XGB** | 63 | 25 | 1 | 0 | 0.99 | 0.99 | 0.98 | 0.99 | 63 | 26 | 0 | 0 | 1.00 | 1.00 | 1.00 | 1.00 |
|  | **MLP** | 61 | 25 | 1 | 2 | 0.97 | 0.95 | 0.96 | 0.96 | 63 | 26 | 0 | 0 | 1.00 | 1.00 | 1.00 | 1.00 |
|  | **SVM** | 62 | 25 | 1 | 1 | 0.98 | 0.97 | 0.97 | 0.97 | 62 | 26 | 0 | 1 | 0.99 | 0.98 | 0.99 | 0.99 |

### Clustering validates the importance of the selected biomolecular signature

Unsupervised clustering was performed by using K-means clustering to further validate the importance of HNSC and OSCC biomolecular signatures. A clear demarcation of two clusters corresponding to two classes, normal and tumor was observed for HNSC and OSCC. It was also noted that clustering based on HNSC and OSCC BioSign data showed a clearer separation than clustering with HNSC and OSCC Top 100 data. The computed SC was between 0.75 – 0.79 whilst high ARI values ranging from 0.70 – 0.91 were attained for all the four clustering tasks. Although correctly clustered HNSC samples decreased from 97.34 % for biomolecular signature to 96.63 % with respect to top 100 features, biomolecular signature candidates were still able to classify in accordance with top 100 features. On the other hand, clustering of OSCC data for features with biomolecular signature improved resulting in a 1% hike for samples being correctly clustered than that of 100 ANOVA features. This finding confirmed that the selected features from OSCC bimolecular signature classifies more precisely the normal and tumor classes than the top 100 features. Unsupervised clustering verified that the selected HNSC and OSCC features from biomolecular signatures yields a clear demarcation of the two classes, normal and tumor samples by performing on par with top 100 features.

### Literature revealing the roles of HNSC and OSCC biomolecular signature candidates in cancer

A literature-based search was performed as an additional validation to confirm the roles of the selected biomarker signatures in HNSC and OSCC. Based on the previous mention of the selected biomolecular signature candidates in the literature, these candidates can be categorised as (i) potential prognostic biomarker and therapeutic candidates, (ii) differentially expressed candidates, and (iii) novel candidates. Tables 3 and 4, respectively list the roles of the candidates selected as HNSC and OSCC biomarker signatures in various cancers including HNSC and OSCC.

**Table 3.** Literature-based roles of selected candidates from HNSC biomolecular signature.

| Category | Biomarker candidate | Cancer | Implication | Reference |
| --- | --- | --- | --- | --- |
| Potential therapeutic candidates | RRAGD, PPL, AQP7, SORT1 | HCC, OV, breast cancer | Roles in inhibiting tumor cell growth, proliferation or metastasis | (Dai et al. 2020; Ding and Liang 2021; Gao et al. 2022; Liu et al. 2022) |
| Potential Prognostic biomarker candidates | GPD1, FAM3D, SASH1, ENDOU, FAM107A, CXCR2, RHCG, SPNS2, PADI1 | Colon cancer, bladder cancer, lung cancer, HNSC, neuroblastoma, fibrosarcoma, RCC, OSCC, ESCC, CRC, LSCC, pancreatic cancer | Tumor suppressor or roles in tumor cell proliferation, migration and invasion | (Qian et al. 2014; Salyakina and Tsinoremas 2016; Ming et al. 2018; Burgess et al. 2020; Xu et al. 2021; Lv et al. 2021; Zhang et al. 2022; Zhu et al. 2023) |
| DEGs | FAM3D, RRAGD, ENDOU, FAM107A, AIF1L | HNSC, OSCC, TSCC, RCC | Down-regulated | (Zhang et al. 2020; Sun et al. 2020; Yang et al. 2021; Fu et al. 2022) |
| DE-miRNAs | PAIP2B | OSCC | Down-regulated | (Crimi et al. 2020) |
| DE-lncRNA | LINC02487, AC091563.1 | HNSC, LUSC | Prognostic biomarker for HNSC, down-regulated | (Li et al. 2020; Guo et al. 2022) |
| Novel candidates | AC091563.1, SMIM5, GBP6 | HNSC, OSCC | SMIM5 may have role in OSCC tumorigenesis while GBP6 and AC091563.1 were down-regulated | (Li et al. 2020; Guo et al. 2022) |
|  | AGFG2, AL356867.1, Z97653.2 | ND | ND | Not reported earlier |

Legends: OV - ovarian cancer, RCC - renal cell carcinoma, ESCC - Esophageal squamous cell carcinoma, CRC - colorectal cancer, LSCC - laryngeal squamous cell carcinoma, LUSC - lung squamous cell carcinoma.

**Table 4.** Literature-based roles of selected candidates from OSCC biomolecular signature.

| Category | Biomarker candidate | Cancer | Implication | Reference |
| --- | --- | --- | --- | --- |
| Potential therapeutic candidates | MAB21L4, RRAGD, UBL3 | cSCC, HCC, lung cancer | Tumor supressor and may have role in cancer proliferation, migration, invasion | (Zhao et al. 2020; Ding and Liang 2021; Srivastava et al. 2022) |
| Potential Prognostic biomarker candidates | ALDH6A1, CAB39L, CEACAM1, EMP1, ENDOU, FHIT, GMDS, GPX3, MUC21, PAX9, SPRR3 | bladder cancer, GC, HNSC, OSCC, lung Adenocarcinoma, ESCC and cancers | Tumor suppressor or tumor promoting roles | (Pickering et al. 2013; Abdalla et al. 2017; Xiong et al. 2018; Wei et al. 2018; Tam et al. 2018; Li et al. 2018; Yoshimoto et al. 2019; Chang et al. 2020; Xu et al. 2021; Fontana et al. 2021; Xia et al. 2023) |
| DEGs | CAB39L, CEACAM1, COBL, DLG2, ENDOU, GPX3, PAX9, PLCXD3, RRAGD, TIMP4 | HNSC, TSCC, OSCC | Down-regulated | (Pedro et al. 2018; Zhang et al. 2020; Sun et al. 2020; Yang et al. 2021; Fu et al. 2022) |
| DE-lncRNA | HCG22 | OSCC | Inhibited cell proliferation, invasion and migration by targeting miR-425-5p | (Fu et al. 2022) |
| Novel candidates | NRG2, SMIM5, MAB21L4 | NSCLC, OSCC, cSCC | May have role in tumorigenesis | (Zhang et al. 2018; Ou et al. 2021) |
|  | TNMD, ZCWPW2 | ND | ND | Not reported earlier |

Legends: cSCC - cutaneous SCC, GC - gastric cancer, NSCLC - non-small cell lung cancer, ESCC - Esophageal squamous cell carcinoma.

#### Potential diagnostic, prognostic and therapeutic biomarkers

Previous studies recommend that nine candidates from HNSC biomolecular signature and eleven candidates from OSCC biomolecular signature have diagnostic and prognostic roles in various cancers (Tables 3 and 4). The findings based on these studies suggested that their involvement in tumor growth, proliferation and metastasis is inevitable and therefore, have been reported as probable diagnostic or prognostic biomarkers. Similarly, the HNSC and OSCC biomarker candidates namely, RRAGD, PPL, AQP7, SORT1, MAB21L4 and UBL3 were earlier considered as potential therapeutic targets due to their regulatory roles in various cancers. These candidates are believed to inhibit cancer proliferation and metastasis by acting as tumor suppressor genes in cancers such as HCC, ovarian cancer (OV), breast cancer (BC), cutaneous squamous cell carcinoma (cSCC) and lung cancer.

#### Differentially expressed candidates within the biomolecular signatures

Other than the candidates with suggested diagnostic, prognostic or therapeutic roles, differentially expressed (DE) candidates such as DEGs and DE-lncRNAs may also be targeted as biomarkers for HNSC and OSCC. 19 HNSC features and 22 OSCC features from the biomolecular signatures were found to be overlapping with DEGs from Shaikh et al. 2019 while few other studies also revealed that the current biomolecular signatures consisted of five HNSC genes and ten OSCC genes that were differentially expressed in various cancers such as HNSC, OSCC, tongue squamous cell carcinoma (TSCC) and renal cell carcinoma (RCC). Likewise, DE-mi-RNA analysis showed that PAIP2B was down-regulated. Likewise, DE-lncRNA analysis presented LINC02487, AC091563.1 and HCG22 to be down-regulated lncRNAs in HNSC, LUSC and OSCC (Li et al. 2020; Fu et al. 2022; Guo et al. 2022).

#### Novel candidates constituting the biomolecular signatures

Nevertheless, roles of novel candidates in HNSC and OSCC are indispensable, and the present study identified novel candidates that are significant and can have crucial roles as biomarkers in HNSC or OSCC. Apart from SMIM5 which has been discussed earlier, a molecular feature of non-small cell lung cancer, NRG2 (Trombetta et al. 2021) was reported in increased levels in breast tumors (Kobayashi et al. 2013). It is also interesting to note that MAB21L4, among the ten significant genes not detected in previous DEG analysis, could potentially have roles in OSCC or HNSC. MAB21L4 was shown to have an essential function in suppressing carcinogenesis for epithelial squamous cell carcinomas (ESCC) (Srivastava et al. 2022) (Srivastava et al. 2022). However, other candidates such as AGFG2, AL356867.1, Z97653.2, TNMD and ZCWPW2 do not have a previous mention in the literature, which opens new avenue to study the role of such novel candidates in cancer.

## Discussion

In the era of next-generation sequencing, volumes of omics data for various cancer sub-types are available. With the advent of machine learning as a data-driven approach, it is now possible to analyse the available omics data and infer meaningful insights towards cancer. The current study focused on developing the ML-based approach to delineate the molecular signatures for HNSC and OSCC. A two-step feature selection strategy, combining ANOVA and SVM-RFE in the subsequent steps, enabled the identification of molecular signatures with respect to HNSC and OSCC. Out of the 100 top features from ANOVA, 90 features in HNSC and 93 features in OSCC were detected as DEGs in a previous study (Shaikh et al. 2019)(Shaikh et al. 2019). This shows that the first step of feature selection, ANOVA eventually captured the 100 statistically significant features in both HNSC and OSCC. The second step of feature selection yielded 22 and 23 significant features, respectively in case of HNSC and OSCC. ML models were trained on six different classifiers utilizing BioSign and Top 100 data pertaining to both HNSC and OSCC. The selected candidates from biomolecular signatures were able to perform well on various classifiers by accurately predicting the tumor versus normal samples. It was found that high classification as well as prediction accuracies can be achieved with the selected features using all the classifiers, confirming that these features are efficient to classify normal and tumor classes for HNSC and OSCC. In addition to increasing the classification accuracy, the biomolecular signatures also made it possible to reduce the number of tumor samples being falsely predicted as normal (false negatives). To further validate the importance of the biomolecular signatures, K-means clustering was performed on BioSign and Top 100 data for both HNSC and OSCC and the results showed a clear demarcation of normal and tumor classes for BioSign data than the respective top 100 data. Together with clustering, literature supported the paramountcy of both the HNSC and OSCC biomolecular signatures, thereby confirming that the selected features can be ideal prognostic biomarkers or therapeutic candidates for HNSC or OSCC.

The selected biomolecular signature candidates fall into the categories of (i) potential prognostic biomarker and therapeutic candidates, (ii) differentially expressed candidates, or (iii) novel candidates. Thirteen HNSC and fourteen OSCC biomolecular signature candidates have been previously identified as potential prognostic biomarker and therapeutic targets for various cancers. Remarkably, a probable biomarker, FAM107A is downregulated during tumors and is associated with several other cancers, such as neuroblastoma, RCC, and fibrosarcoma (Salyakina and Tsinoremas 2016; Sun et al. 2020). Similarly, previous studies showed that FAM3D has been shown to be one among the down-regulated genes during DEG analysis of HNSC, and its expression in patients’ blood was proposed as an early biomarker for colon cancer (Salyakina and Tsinoremas 2016; Sun et al. 2020). Another significant gene, CXCR2, being a receptor of CXC chemokines, plays a critical role in the invasion and metastases of OSCC (Qian et al. 2014). The experimental data of CXCR2 and its existence on normal as well as cancerous oral epithelial cells suggested its role clearly in oral cancer biology as biomarker (Khurram et al. 2014)

Differentially expressed candidates such as DEGs and DE-lncRNAs are also critical and contribute significantly towards cancer progression and metastasis. The gene CAB39L which was downregulated during DEG analysis of OSCC transcriptomics data (Lo et al. 2012; Yang et al. 2021, Wan et al. 2022) showed to express in higher amounts in the tumor tissues from the same patients (Lo et al. 2012). Other than above-mentioned genes, DLG2, PLCXD3, and TIMP4 have been found to be down-regulated genes based on the DEG analysis of HNSC by Sun et al. 2020Sun *et al*. 2020. Nevertheless, the association of long noncoding RNA (lncRNA) in cancers particularly HNSC and OSCC have been discussed earlier (Ghosh and Majumder 2022) and shown to be important by means of differential expression analysis of lncRNA (Li et al. 2019). LINC02487 and AC091563.1 were earlier reported as DE-lncRNA for HNSC (Li et al. 2019), while other studies revealed that LINC02487 can act as a tumor suppressor through the USP17–SNAI1 axis (Feng et al. 2020) and that AC091563.1 has also a role as a DE-lncRNA in case of TSCC (Zhang et al. 2020). Interestingly, HCG22, another important lncRNA related to plasma membrane, is usually downregulated during OSCC but experiments revealed that increased HCG22 expression tend to inhibit the cellular proliferation of OSCC (Fu et al. 2022). The present study included a list of novel HNSC and OSCC biomarker candidates that have not been reported previously or with not known functions. However, these novel candidates are also found to be significant in HNSC and OSCC which opens up a new avenue to study the role of such candidates as prognostic biomarker or therapeutic agent in cancer. For instance, one such candidate, MAB21L4, was not detected as a DEG during previous analyses but more likely suggests it’s potential role in OSCC or HNSC due to it’s essential function in suppressing carcinogenesis for ESCC (Srivastava et al. 2022). The current ML pipeline successfully identified the HNSC and OSCC biomolecular signatures consisting of biomarkers, differentially expressed and novel candidates that have significant roles in HNSC, OSCC or other cancers and could potentially be targeted for diagnostic, prognostic or therapeutic purposes.

Specifically, out of the 22 HNSC and 23 OSCC biomolecular signature candidates, three genes namely, ENDOU, RRAGD, and SMIM5 were the overlapping genes among the significant features for both the cancers, HNSC and OSCC and therefore, can be used as a common biomarker for HNSC and OSCC. RRAGD is also known to be a critical factor in cancer progression and potentially acts as a therapeutic target during human hepatocellular carcinoma (HCC) (Ding and Liang 2021). It is also identified as a down-regulated gene during DEG analysis of TSCC and OSCC (Zhang et al. 2020). Similarly, ENDOU, a down-regulated gene in OSCC, has also indicated its role as a tumor suppressor during HNSC progression (Xu et al. 2021). The third common gene, SMIM5, a new molecule reported only in a few studies regarding its role in OSCC, requires further research to establish its potential role in HNSC or OSCC (Zhang et al. 2018). Nevertheless, there are other HNSC biomarker signature candidates identified in the present study that are overlapping with previously reported significant genes to be important in OSCC and vice versa (Tables 3 and 4) suggesting that these common features could also be targeted as biomarkers for both HNSC and OSCC, together.

Another key finding of the current ML-based analysis is that the biomarker signatures included seven genes, namely, CEACAM1, COBL, GPX3, HCG22, MUC21, PAX9 and SMIM5, that are associated with the cellular or plasma membrane, making these features potential candidate for non-invasive methods of bio-diagnostics in HNSC and OSCC. The hypothesis is supported by previous individual studies where roles of these features are prominent in cancer. For instance, high amounts of CEACAM1 are reported in oral cancer, and it is shown that CEACAM1 is related to tumorigenesis (Wang et al. 2017). It has also been concluded that targeting CEACAM1 can yield a novel immune-therapeutical biomarker for HNSC (Tam et al. 2018). DEG analysis of micro-array data revealed that CEACAM1 and COBL are amongst the total 261 DEGs during OSCC (Yang et al. 2021). Likewise, GPX3 was found to be downregulated in many cancers which may lead to cancer progression and therefore act as a potential prognostic biomarker (Pedro et al. 2018; Chang et al. 2020). Another study study revealed that higher HCG22 levels prevented the invasion and migration of cells during OSCC via regulating the expression of the miRNA, miR-425-5p (Fu et al. 2022) while Yoshimoto et al. 2019 showed that MUC21, expressed on cell surface, is involved in cell incohesiveness in lung adrenocarcinoma. It is also, noteworthy to mention PAX9’s involvement in regulating squamous cell differentiation and carcinogenesis in ESCC (Xiong et al. 2018) and as a tumor suppressor gene in various other cancers (Bhol et al. 2022) that makes it an ideal candidate for as a therapeutic target. Similarly, it has been reported earlier that one of the common genes from HNSC and OSCC biomolecular signatures, SMIM5 is also involved in the OSCC tumorogenesis (Zhang et al. 2018). The seven candidates may prove to be excellent candidates as potential biomarkers due to their relation with plasma membrane making it easier to target for diagnostics, prognostic or therapeutic purposes.

### Concluding remarks

The present study enabled the identification of biomolecular signatures that exhibited their imperative roles in HNSC and OSCC. A two-step feature selection strategy employed to establish the biomolecular signature candidates based on which ML models were trained and tested independently. The results showed that the biomolecular signatures enabled not only accurate classification and prediction but also the clustering of samples into tumor and normal classes. Literature-based search validated the significant roles of the selected features from the HNSC and OSCC biomolecular signatures in cancer. The biomolecular signature candidates, in particular the three overlapping genes and seven plasma-membrane related features, could serve as potential diagnostic or prognostic or therapeutic candidates.

## Data Availability

All data downloaded from Genomic Data Commons (GDC) Data Portal (https://portal.gdc.cancer.gov/)

